# An exploratory machine learning study on paediatric abdominal pain phenotyping and prediction

**DOI:** 10.1101/2023.04.26.23289185

**Authors:** Kazuya Takahashi, Michalina Lubiatowska, Huma Shehwana, James K. Ruffle, John A Williams, Animesh Acharjee, Shuji Terai, Georgios V Gkoutos, Humayoon Satti, Qasim Aziz

**Affiliations:** Centre for Neuroscience and Trauma, Wingate Institute of Neurogastroenterology, Blizard Institute, Barts and The London School of Medicine and Dentistry, Queen Mary University of London, London, UK; Division of Gastroenterology and Hepatology, Graduate School of Medical and Dental Sciences, Niigata University, Niigata, Japan; Department of Biological Sciences, National University of Medical Sciences, Rawalpindi, Pakistan; Queen Square Institute of Neurology, University College London, UK; College of Medical and Dental Sciences, Institute of Cancer and Genomic Sciences, University of Birmingham, Birmingham, UK; Health Data Research UK, Midlands Site, Birmingham, UK; Centre for Health Data Science, Birmingham, UK

**Keywords:** paediatric abdominal pain, machine learning, artificial intelligence, Uniform Manifold Approximation and Projection, Shapley Additive exPlanations

## Abstract

**Background:** The exact mechanisms underlying paediatric abdominal pain (AP) remain unclear due to patient heterogeneity. This study aimed to identify AP phenotypes and develop predictive models to explore associated factors.

**Methods:** In 13,790 children from a large birth cohort, data on paediatric and maternal demographics and comorbidities were extracted from general practitioner records. Machine learning (ML) clustering was used to identify distinct AP phenotypes, and an ML-based predictive model was developed using demographics and clinical features.

**Results:** 1,274 children experienced AP (9.2 %) (average age: 8.4 ± 1.1 years, male/female: 615/659), who clustered into three distinct phenotypes: Phenotype 1 with an allergic predisposition (n = 137), Phenotype 2 with maternal comorbidities (n = 676), and Phenotype 3 with minimal other comorbidities (n = 340). As the number of allergic diseases or maternal comorbidities increased, so did the frequency of AP, with 17.6% of children with ≥ 3 allergic diseases and 25.6% of children with ≥ 3 maternal comorbidities. The predictive model demonstrated moderate performance in predicting paediatric AP (AUC 0.67), showing that a child’s ethnicity, paediatric allergic diseases, and maternal comorbidities were key predictive factors. When stratified by ML-predicted probability, observed AP rates were 18.9% in the <40% group, 44.8% in the 40–50% group, 60.6% in the 50–60% group, and 100.0% in the >60% group.

**Conclusions:** Our findings reveal distinct phenotypes and associated factors of paediatric AP by an ML approach. These insights suggest potential targets for future research to clarify the underlying mechanisms of paediatric AP.

## 1 Introduction

Abdominal pain (AP) is one of the most common symptoms among children and adolescents, with prevalence rates across the USA and Europe ranging from 0.3% to 19.0% [1–3]. In primary care, children presenting with AP are diagnosed with functional or medically un-explained AP in 80% of the cases, while organic causes are considerably less frequent [4,5]. Although rarely life-threatening, AP is often refractory to treatment and associated with psychiatric comorbidities, such as anxiety and depressive disorders [6], significantly affecting health-related quality of life [7]. Therefore, early recognition and intervention are warranted.

Organic diseases, as well as early life events ranging from allergy status, psychological comorbidity, and parental factors, are proposed risk factors for paediatric AP [5,8–10]. Moreover, these diverse risk factors can interact, contributing to its complexity and heterogeneity. A more robust stratification with large patient cohorts is essential for understanding the aetiology of paediatric AP, plausibly disclosing novel insights into its underlying mechanisms.

A robust approach for identifying subgroups of patients with shared characteristics is data-driven clustering by unsupervised machine learning (ML) [11,12]. Any yielded subgroups may share an underlying mechanism associated with AP. Furthermore, supervised ML is considered a powerful tool for clinical outcome prediction [13], and it could aid clinicians in assessing the risk of AP development in early childhood [14]. Based on this background, we hypothesised that an ML algorithm could classify children with AP into distinct phenotypes based on common characteristics, thereby helping to unravel the complex underlying AP mechanisms. Additionally, we hypothesised that ML algorithms could predict the development of paediatric AP and help identify the key factors associated with it.

In this exploratory study, we comprehensively evaluated the risk factors of paediatric patients with AP in a large birth cohort, covering a broad spectrum of cases. Using unsupervised ML, we delineated phenotypes of paediatric AP using paediatric and maternal clinical data. Moreover, ML models tasked to predict the development of paediatric AP uncovered underlying factors linked to its frequency, catalytic for future hypothesis-driven research.

## 2 Materials and Methods

### 2.1 Participants

Between 2007 and 2011, 12,453 pregnant women (recruited at 26–28 weeks) and 13,858 children were registered in the Born in Bradford (BiB) cohort. The detailed demographics of the whole cohort are described elsewhere [15]. Data for this study were obtained from the BiB cohort under agreement, with informed consent collected at the time of data gathering. For this study, we included children who had linked general practitioner (GP) records and whose comorbidities could be identified using the systematized nomenclature of medicine clinical terms (SNOMED-CT), a standardized terminology system used in medicine and healthcare. The details of the study population are illustrated in Fig 1. Ethical approval was obtained from the Bradford Research Ethics Committee (Ref 07/H1302/112). This study was conducted following the principles of the Declaration of Helsinki.

**FIGURE 1.**
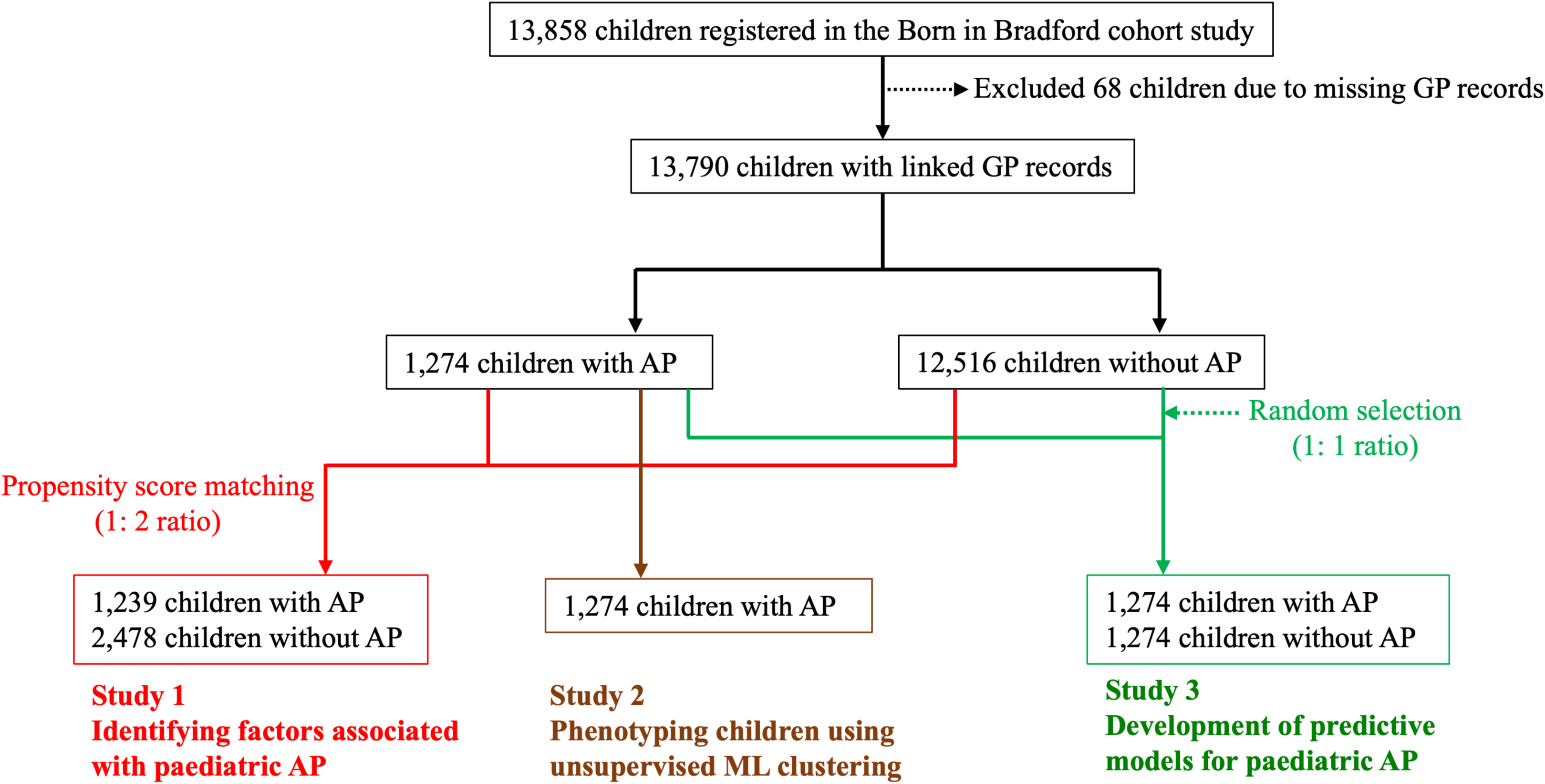
Study population. Among 13,858 children in the Born in Bradford cohort, 13,790 with linked general practitioner (GP) records were included. Study 1 identified factors associated with paediatric abdominal pain (AP) using propensity score matching (1:2 ratio). Study 2 performed phenotyping of AP using unsupervised machine learning (ML) clustering. Study 3 developed predictive models using supervised ML, including 1,274 children with AP and 1,274 randomly selected children without AP (1:1 ratio).

### 2.2 Definition of AP and frequency survey of paediatric and maternal comorbidities

AP was defined as the presence of one of the following diagnoses in SNOMED-CT, regardless of its cause or whether it was acute or chronic: AP, abdominal wall pain, or generalized AP. The frequencies of comorbidities, such as gastrointestinal (GI), psychological, and allergic diseases (asthma, eczema, urticaria, or hay fever), as well as diseases causing somatic pain, were also retrieved using SNOMED-CT, since these are associated with both organic and functional AP [8,10,16–18]. Additionally, the comorbidities of the children’s genetic mothers were extracted to identify any associations between the maternal comorbidities and paediatric AP. The diseases of the children and their mothers investigated in this study and their SNOMED-CT codes are listed in S1 Table. Fathers were not included in our analysis pipeline due to a significant proportion of missing values.

### 2.3 Study 1. Identifying factors associated with paediatric AP

Study 1 aimed to determine the frequency of AP, elucidate the clinical characteristics of children with AP, and identify its associated factors. First, to adjust for background differences at the time of extraction between children with and without AP, we employed propensity score matching. After matching, we compared the two matched groups to assess differences in demographics and the frequency of comorbidities.

Next, we identified factors associated with paediatric AP. For this analysis, all comorbidities listed in S1 Table were included, irrespective of whether they occurred before or after the onset of AP, as the aim was to examine their overall association with paediatric AP.

### 2.4 Study 2. Phenotyping paediatric AP using unsupervised ML clustering

Study 2 focused on children experiencing AP, using unsupervised ML clustering to delineate phenotypes. In Python (version 3.7.12) [19], Uniform Manifold Approximation and Projection (UMAP), a non-linear dimensionality reduction technique, was applied to embed all variables into a three-dimensional latent space for subsequent clustering [20]. A scatterplot was then generated from the latent features, and phenotypic AP clusters were identified using Hierarchical Density-based Spatial Clustering of Applications with Noise (HDBSCAN), a density-based hierarchical method [21]. Demographics and comorbidities of both children and their mothers were subsequently compared across the identified phenotypes to elucidate their distinct characteristics. Furthermore, we investigated whether the number of significant comorbidities identified in the phenotype analysis was associated with the frequency of paediatric AP.

### 2.5 Study 3. Using supervised ML to predict the development of paediatric AP

To construct a balanced dataset, we randomly selected an equal number of children without AP from the control group to match those with AP (Fig 1), thereby mitigating potential model bias due to class imbalance. Predictive variables included demographic information and pre-existing comorbidities diagnosed prior to the onset of AP. Comorbidities with a prevalence of less than 1% were excluded to reduce the risk of model overfitting. The variables used in the predictive model, along with their frequencies in children with and without AP, are summarised in S2 Table. The overall workflow for model development is illustrated in S1 Fig.

We initially developed predictive models for paediatric AP using several ML algorithms, including eXtreme Gradient Boosting, Categorical Boosting (CatBoost), Light Gradient Boosting Machine, and Random Forest. Model performance was assessed using receiver operating characteristic (ROC) curves and the corresponding area under the curve (AUC). In addition, accuracy, sensitivity, specificity, positive predictive value (PPV), and negative predictive value (NPV) were calculated. The model with the highest AUC was selected for subsequent feature importance analysis and risk stratification.

To interpret model predictions, we employed Shapley Additive exPlanations (SHAP) values. SHAP quantifies the contribution of each feature to individual predictions, allowing us to identify which variables had the greatest influence on the model’s outputs and to enhance interpretability [22,23].

Finally, because the ML models produce continuous probability estimates for AP, we stratified children in the test dataset into four probability groups: <40%, 40–50%, 50– 60%, and >60%. The observed risk of paediatric AP was then compared across these strata.

### 2.6 Statistical analysis

Continuous data were expressed as mean ± standard deviation (SD). Categorical data were expressed as numbers and percentages. The Student’s t-test and Chi-squared test were used for numerical and categorical data, respectively.

To compare children with AP and those without, the propensity score was calculated using logistic regression analysis, with age at extraction and gender as covariates. Matched children with and without AP were then extracted. A 1:2 nearest-neighbour matching without replacement was performed, with a calliper width set to 0.2 times the standard deviation of the logit of the propensity score. Model discrimination was assessed using the c-statistic, and covariate balance was evaluated using standardized mean differences (SMDs), with SMD < 0.1 indicating good balance.

Univariate and multivariate logistic regression analyses were utilised to calculate odds ratios (ORs) and 95% confidence intervals (CI) for each variable, determining variables associated with paediatric AP. Variables significant in the univariate analysis were included in the multivariate analysis. As the comparisons between phenotypes were descriptive and intended to characterize group differences, Bonferroni correction was not applied. The Wald test was applied to compare ORs among the predicted probability groups. A p-value < 0.05 was considered statistically significant. EZR (Saitama Medical Center, Jichi Medical University, Saitama, Japan) was used for statistical analysis [24].

## 3 Results

### 3.1 Study 1. Identifying factors associated with paediatric AP

A total of 13,790 children were included in the analysis, of whom 1,274 (9.2%) experienced AP (male/female = 615/659). The average age at diagnosis of AP was 5.6 ± 2.7 years. The characteristics of all children are summarised in S3 Table.

As a result of propensity score matching, 1,239 children with AP and 2,478 children without AP were selected (Table 1). Compared to children without AP, more children of Pakistani origin were in the AP group, while fewer were of White origin (all *p* < 0.01). Children with AP had higher rates of comorbidities, including allergic diseases, appendicitis, Celiac disease, constipation, and gastro-oesophageal reflux disease (GORD), and migraine (all *p* < 0.01). Moreover, mothers of children with AP reported higher incidences of AP, allergic diseases, arthritis, chronic muscle pain, functional dyspepsia, GORD, irritable bowel syndrome (IBS), and migraine (all *p* < 0.01).

**Table 1.** Comparison of children with and without abdominal pain.

|  | Before matching (n = 13,790) |  |  | After matching (n = 3,717) |  |  |
| --- | --- | --- | --- | --- | --- | --- |
|  | Without AP<br>(n=12,516) | With AP<br>(n=1,274) | *p* value | Without AP<br>(n=2,478) | With AP<br>(n=1,239) | *p* value |
| Average age at the extraction ± SD<br>(years) | 8.3 ± 1.1 | 8.4 ± 1.1 | <0.01 | 8.6 ± 1.1 | 8.6 ± 1.1 | 0.83 |
| Average age at the diagnosis of<br>abdominal pain ± SD (years) | N/A | 5.6 ± 2.7 | N/A | N/A | 7.5 ± 2.3 | N/A |
| Gender, female, n (%) | 6,015 (48.1 %) | 659 (51.7) | 0.01 | 1,304 (52.6) | 634 (51.2) | 0.40 |
| Route of birth, (vaginal), n (%) | 9,415 (77.0 %) | 943 (75.3) | 0.17 | 1,866 (75.3) | 916 (73.9) | 0.36 |
| Ethnicity*, n (%) |  |  |  |  |  |  |
| Pakistani | 4,750 (45.8) | 703 (58.6) | <0.01 | 970 (46.9) | 681 (58.4) | <0.01 |
| White | 3,771 (36.4) | 267 (22.3) | <0.01 | 724 (35.0) | 259 (22.2) | <0.01 |
| Other ethnicities | 1,847 (17.8) | 229 (19.1) | 0.27 | 374 (18.1) | 227 (19.5) | 0.34 |
| **Frequency of diseases, n (%)** |  |  |  |  |  |  |
| Allergic diseases | 4,665 (37.3) | 601 (47.2) | <0.01 | 918 (37.0) | 584 (47.1) | <0.01 |
| Appendicitis | 24 (0.2) | 27 (2.1) | <0.01 | 4 (0.2) | 27 (2.2) | <0.01 |
| Arthritis | 6 (0.0) | 1 (0.1) | 0.49 | 0 (0.0) | 1 (0.1) | 0.16 |
| Celiac disease | 24 (0.2) | 9 (0.7) | <0.01 | 3 (0.1) | 9 (0.7) | <0.01 |
| Constipation | 142 (1.1) | 57 (4.5) | <0.01 | 30 (1.2) | 56 (4.5) | <0.01 |
| FD | 3 (0.0) | 4 (0.3) | <0.01 | 2 (0.1) | 4 (0.3) | 0.08 |
| GORD | 342 (2.7) | 66 (5.2) | <0.01 | 54 (2.2) | 63 (5.1) | <0.01 |
| IBD, colitis | 4 (0.0) | 2 (0.2) | 0.1 | 1 (0.0) | 2 (0.2) | 0.22 |
| Migraine | 53 (0.4) | 16 (1.3) | <0.01 | 10 (0.4) | 16 (1.3) | <0.01 |
| EDS, JHS | 9 (0.1) | 4 (0.3) | <0.01 | 1 (0.0) | 4 (0.3) | 0.03 |
| Autism | 36 (0.3) | 4 (0.3) | 0.87 | 9 (0.4) | 4 (0.3) | 0.84 |
| Intellectual disability | 16 (0.1) | 2 (0.2) | 0.78 | 3 (0.1) | 2 (0.2) | 0.75 |
| Mother's abdominal pain | 3,985 (31.8) | 631 (49.5) | <0.01 | 782 (31.6) | 616 (49.7) | <0.01 |
| Mother's allergic disease | 4,931 (39.4) | 609 (47.8) | <0.01 | 965 (38.9) | 597 (48.2) | <0.01 |
| Mother's appendicitis | 94 (0.8) | 13 (1.0) | 0.31 | 14 (0.6) | 13 (1.0) | 0.10 |
| Mother's arthritis | 495 (4.0) | 109 (8.6) | <0.01 | 24 (1.0) | 28 (2.3) | <0.01 |
| Mother's Celiac disease | 73 (0.6) | 14 (1.1) | 0.04 | 17 (0.7) | 13 (1.0) | 0.24 |
| Mother's chronic fatigue syndrome | 25 (0.2) | 3 (0.2) | 0.74 | 6 (0.2) | 3 (0.2) | 1.00 |
| Mother's chronic muscle pain | 167 (1.3) | 30 (2.4) | 0.01 | 30 (1.2) | 29 (2.3) | <0.01 |
| Mother's constipation | 128 (1.0) | 10 (0.8) | 0.55 | 36 (1.5) | 10 (0.8) | 0.09 |
| Mother's depressive disorder, bipolar disorder | 2,632 (21.0) | 309 (24.3) | 0.01 | 555 (22.4) | 300 (24.2) | 0.22 |
| Mother's FD | 162 (1.3) | 33 (2.6) | <0.01 | 36 (1.5) | 33 (2.7) | <0.01 |
| Mother's GORD | 876 (7.0) | 171 (13.4) | <0.01 | 172 (6.9) | 170 (13.7) | <0.01 |
| Mother's IBD | 84 (0.7) | 10 (0.8) | 0.59 | 12 (0.5) | 9 (0.7) | 0.35 |
| Mother's IBS | 877 (7.0) | 126 (9.9) | <0.01 | 171 (6.9) | 125 (10.1) | <0.01 |
| Mother's migraine | 1,845 (14.7) | 285 (22.4) | <0.01 | 378 (15.3) | 283 (22.8) | <0.01 |
| Mother's EDS, JHS | 47 (0.3) | 5 (0.4) | 0.93 | 10 (0.4) | 5 (0.4) | 1.00 |
| Mother's obsessive-compulsive disorder | 61 (0.5) | 10 (0.8) | 0.15 | 13 (0.5) | 10 (0.8) | 0.30 |
| Mother's schizophrenia | 6 (0.0) | 2 (0.2) | 0.17 | 1 (0.0) | 1 (0.1) | 0.62 |
| Mother's intellectual disability | 44 (0.4) | 1 (0.1) | 0.1 | 8 (0.3) | 1 (0.1) | 0.16 |

\* Gender and age at extraction were used to calculate the propensity score. Before matching, the standardized mean differences (SMD) for gender and age were 0.073 and 0.337, respectively. After matching, the SMD values for gender and age were reduced to 0.029 and 0.007, respectively, indicating that the data were well balanced. The c-statistic was 0.60 (95% CI: 0.58–0.61).
AP, abdominal pain; CI, confidence interval; SD, standard deviation; FD, functional dyspepsia; GORD, gastro-oesophageal reflux disease; IBD, inflammatory bowel disease; IBS, irritable bowel syndrome; EDS, Ehlers-Danlos syndrome; JHS, joint hypermobility syndrome

Multivariate logistic regression analysis revealed that Pakistani ethnicity, allergic diseases, GI diseases, migraine, maternal AP, maternal allergic diseases, maternal GORD, and maternal migraine were significantly associated with paediatric AP (Table 2). Non–significant results are summarized in S4 Table.

**Table 2.** Significant results of logistic regression analysis for the diagnosis of paediatric abdominal pain.

|  | Univariate |  | Multivariate |  |
| --- | --- | --- | --- | --- |
|  | OR (95% CI) | *p* value | OR (95% CI) | *p* value |
| Pakistani (vs. other ethnicities) | 1.59 (1.37–1.83) | <0.01 | 1.58 (1.39–1.79) | <0.01 |
| Allergic diseases | 1.52 (1.32–1.74) | <0.01 | 1.19 (1.05–1.34) | 0.01 |
| Appendicitis | 13.8 (4.81–39.5) | <0.01 | 8.92 (4.94–16.1) | <0.01 |
| Celiac disease | 6.04 (1.63–22.3) | 0.01 | 3.35 (1.53–7.34) | <0.01 |
| Constipation | 3.86 (2.47–6.05) | <0.01 | 3.11 (2.23–4.33) | <0.01 |
| GORD | 2.40 (1.66–3.48) | <0.01 | 1.64 (1.23–2.18) | <0.01 |
| Migraine | 3.23 (1.46–7.14) | <0.01 | 2.95 (1.64–5.33) | <0.01 |
| Mother's abdominal pain | 2.14 (1.86–2.47) | <0.01 | 1.68 (1.48–1.90) | <0.01 |
| Mother's allergic diseases | 1.46 (1.27–1.67) | <0.01 | 1.17 (1.03–1.33) | 0.01 |
| Mother's arthritis | 2.36 (1.36–4.10) | <0.01 | 1.39 (0.89–2.16) | 0.14 |
| Mother's chronic fatigue syndrome | 1.96 (1.17–3.27) | 0.01 | 0.98 (0.28–3.41) | 0.98 |
| Mother's chronic muscle pain | 1.96 (1.17–3.27) | 0.01 | 1.12 (0.73–1.72) | 0.61 |
| Mother's FD | 1.86 (1.15–2.99) | 0.01 | 1.15 (0.76–1.74) | 0.51 |
| Mother's GORD | 2.13 (1.70–2.67) | <0.01 | 1.46 (1.21–1.76) | <0.01 |
| Mother's IBS | 1.51 (1.19–1.93) | <0.01 | 1.07 (0.86–1.33) | 0.55 |
| Mother's migraine | 1.64 (1.38–1.95) | <0.01 | 1.24 (1.06–1.44) | 0.01 |

FD, functional dyspepsia; GORD, gastro-oesophageal reflux disease; IBD, inflammatory bowel disease; IBS, irritable bowel syndrome; EDS, Ehlers-Danlos syndrome; JHS, joint hypermobility syndrome

### 3.2 Study 2. Phenotyping children using unsupervised ML clustering

The unsupervised model classified children with AP into three distinct phenotypes: 137 children in Phenotype 1 (10.8%), 677 children in Phenotype 2 (53.1%), and 340 children in Phenotype 3 (26.7%) (Figure 2). The remaining 120 children (9.4%) exhibited miscellaneous characteristics, making them unclassifiable by the model; these children were excluded from downstream analyses.

**FIGURE 2.**
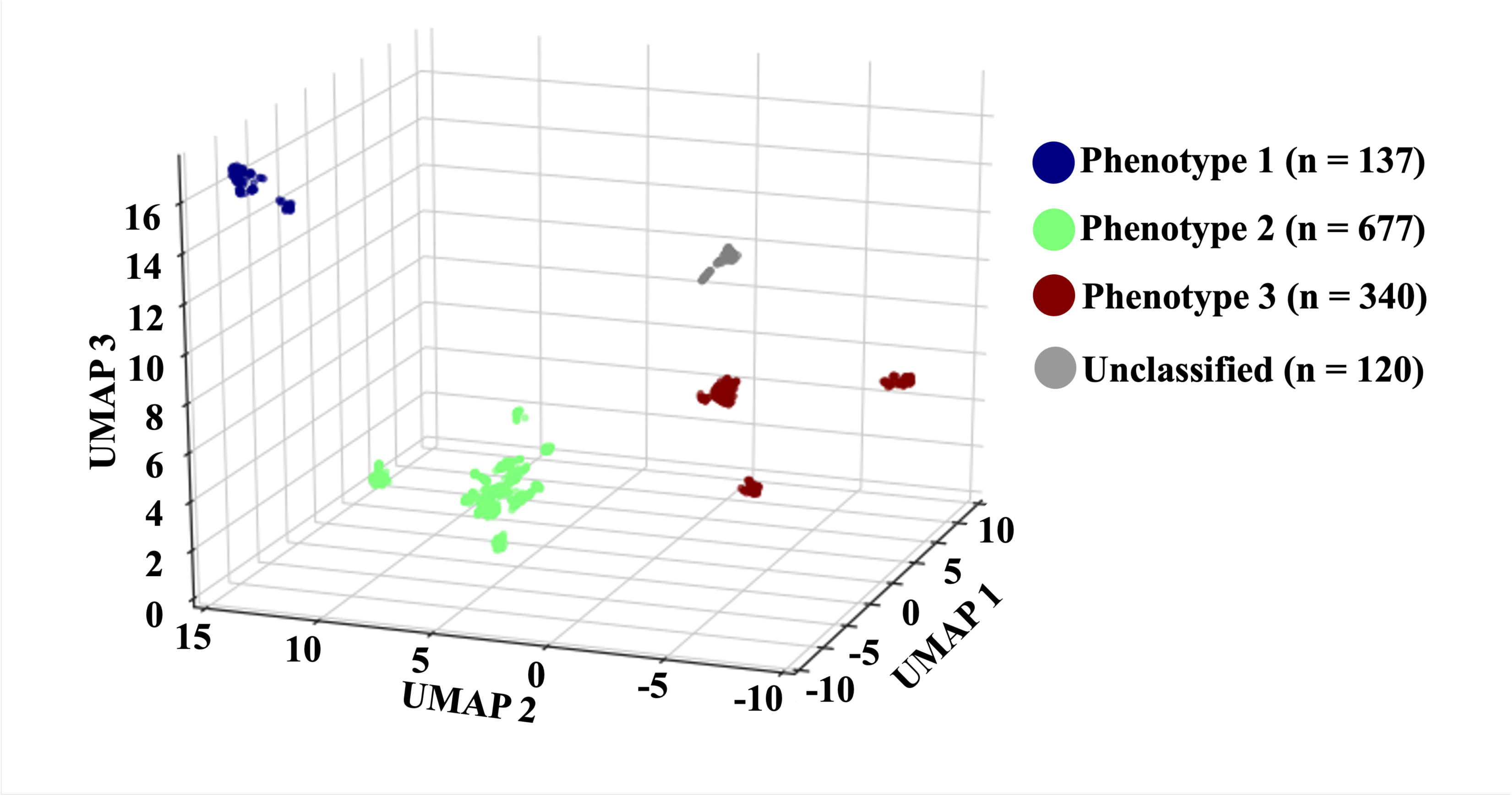
Phenotyping of children with abdominal pain. Scattering plots of based on three new variables created by unsupervised Uniform Manifold Approximation and Projection (UMAP). Hierarchical Density-Based Spatial Clustering of Applications with Noise (HDBSCAN) found 3 distinct phenotypes.

Subsequently, we compared the clinical characteristics among the three phenotypes (Table 3). Age at diagnosis of AP, gender, route of birth, and ethnicity showed almost no significant differences among the three phenotypes. The frequency of GI disorders was low in all phenotypes and showed few significant differences between the phenotypes. The clinical characteristics of each phenotype are summarised below.

**Table 3.**
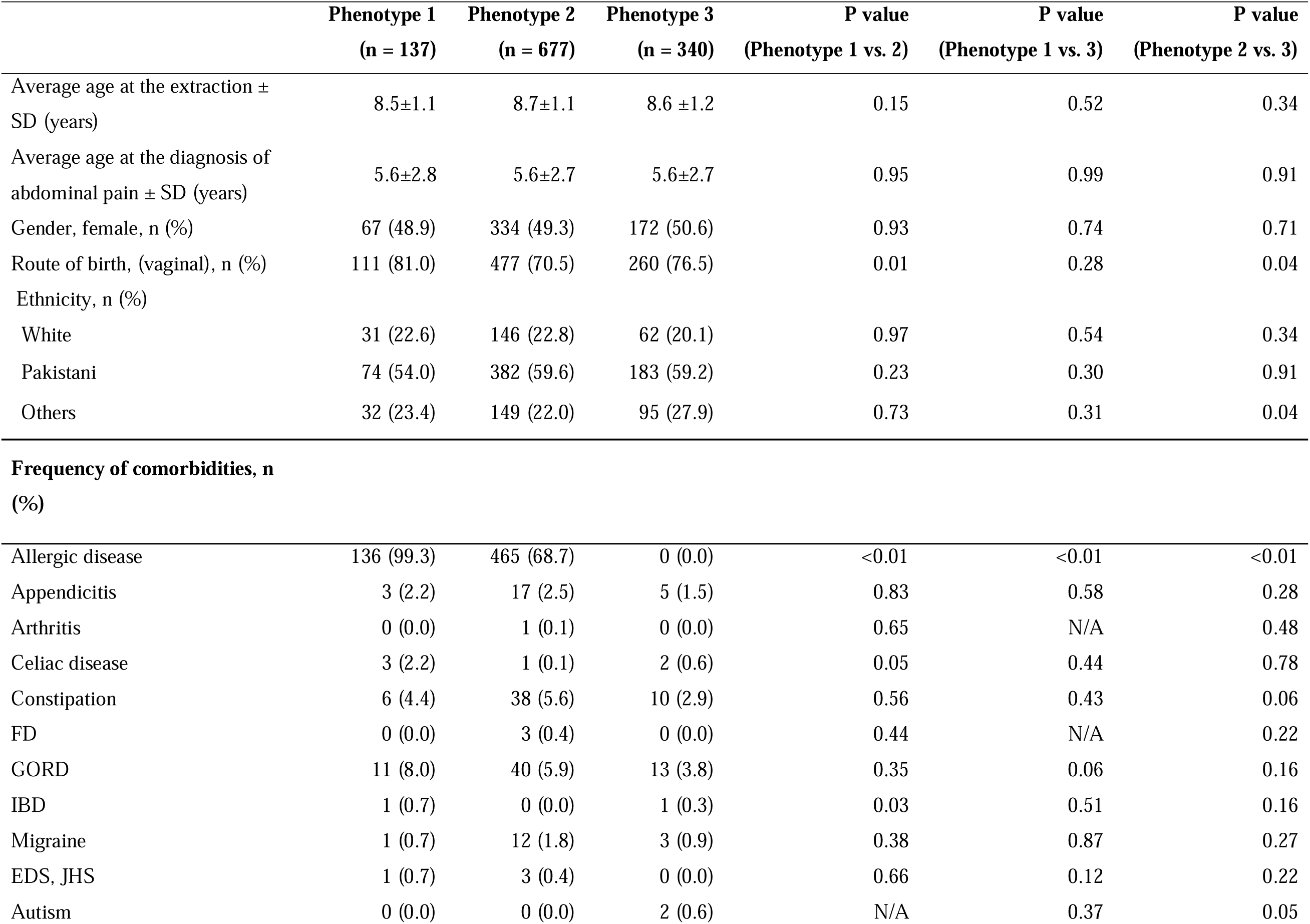
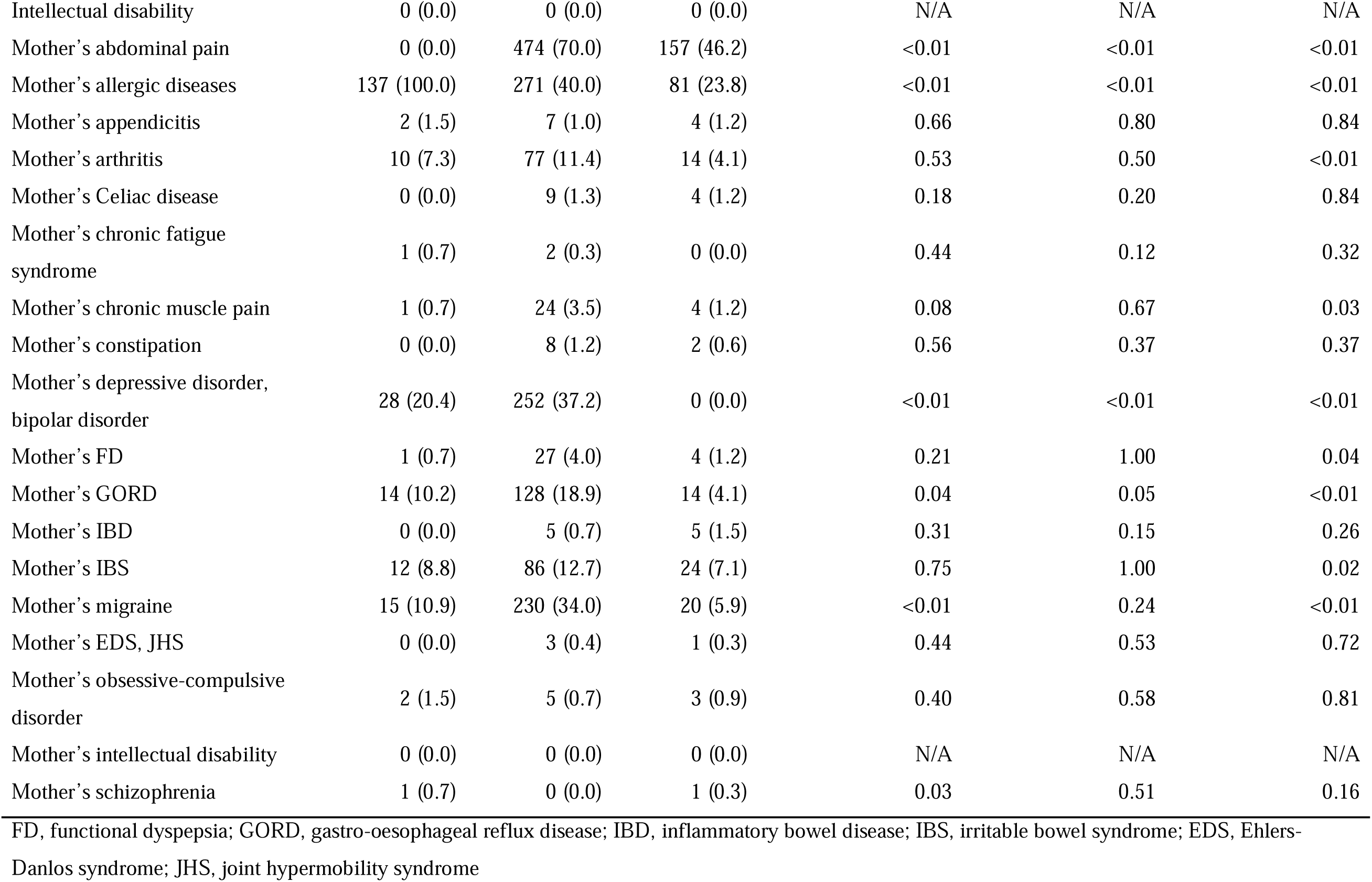
Comparison of clinical characteristics among 3 pain phenotypes.

|  | Phenotype 1<br>(n = 137) | Phenotype 2<br>(n = 677) | Phenotype 3<br>(n = 340) | P value<br>(Phenotype 1 vs. 2) | P value<br>(Phenotype 1 vs. 3) | P value<br>(Phenotype 2 vs. 3) |
| --- | --- | --- | --- | --- | --- | --- |
| Average age at the extraction ± SD (years) | 8.5±1.1 | 8.7±1.1 | 8.6 ±1.2 | 0.15 | 0.52 | 0.34 |
| Average age at the diagnosis of abdominal pain ± SD (years) | 5.6±2.8 | 5.6±2.7 | 5.6±2.7 | 0.95 | 0.99 | 0.91 |
| Gender, female, n (%) | 67 (48.9) | 334 (49.3) | 172 (50.6) | 0.93 | 0.74 | 0.71 |
| Route of birth, (vaginal), n (%) | 111 (81.0) | 477 (70.5) | 260 (76.5) | 0.01 | 0.28 | 0.04 |
| Ethnicity, n (%) |  |  |  |  |  |  |
| White | 31 (22.6) | 146 (22.8) | 62 (20.1) | 0.97 | 0.54 | 0.34 |
| Pakistani | 74 (54.0) | 382 (59.6) | 183 (59.2) | 0.23 | 0.30 | 0.91 |
| Others | 32 (23.4) | 149 (22.0) | 95 (27.9) | 0.73 | 0.31 | 0.04 |
| **Frequency of comorbidities, n (%)** |  |  |  |  |  |  |
| Allergic disease | 136 (99.3) | 465 (68.7) | 0 (0.0) | <0.01 | <0.01 | <0.01 |
| Appendicitis | 3 (2.2) | 17 (2.5) | 5 (1.5) | 0.83 | 0.58 | 0.28 |
| Arthritis | 0 (0.0) | 1 (0.1) | 0 (0.0) | 0.65 | N/A | 0.48 |
| Celiac disease | 3 (2.2) | 1 (0.1) | 2 (0.6) | 0.05 | 0.44 | 0.78 |
| Constipation | 6 (4.4) | 38 (5.6) | 10 (2.9) | 0.56 | 0.43 | 0.06 |
| FD | 0 (0.0) | 3 (0.4) | 0 (0.0) | 0.44 | N/A | 0.22 |
| GORD | 11 (8.0) | 40 (5.9) | 13 (3.8) | 0.35 | 0.06 | 0.16 |
| IBD | 1 (0.7) | 0 (0.0) | 1 (0.3) | 0.03 | 0.51 | 0.16 |
| Migraine | 1 (0.7) | 12 (1.8) | 3 (0.9) | 0.38 | 0.87 | 0.27 |
| EDS, JHS | 1 (0.7) | 3 (0.4) | 0 (0.0) | 0.66 | 0.12 | 0.22 |
| Autism | 0 (0.0) | 0 (0.0) | 2 (0.6) | N/A | 0.37 | 0.05 |
| Intellectual disability | 0 (0.0) | 0 (0.0) | 0 (0.0) | N/A | N/A | N/A |
| Mother's abdominal pain | 0 (0.0) | 474 (70.0) | 157 (46.2) | <0.01 | <0.01 | <0.01 |
| Mother's allergic diseases | 137 (100.0) | 271 (40.0) | 81 (23.8) | <0.01 | <0.01 | <0.01 |
| Mother's appendicitis | 2 (1.5) | 7 (1.0) | 4 (1.2) | 0.66 | 0.80 | 0.84 |
| Mother's arthritis | 10 (7.3) | 77 (11.4) | 14 (4.1) | 0.53 | 0.50 | <0.01 |
| Mother's Celiac disease | 0 (0.0) | 9 (1.3) | 4 (1.2) | 0.18 | 0.20 | 0.84 |
| Mother's chronic fatigue syndrome | 1 (0.7) | 2 (0.3) | 0 (0.0) | 0.44 | 0.12 | 0.32 |
| Mother's chronic muscle pain | 1 (0.7) | 24 (3.5) | 4 (1.2) | 0.08 | 0.67 | 0.03 |
| Mother's constipation | 0 (0.0) | 8 (1.2) | 2 (0.6) | 0.56 | 0.37 | 0.37 |
| Mother's depressive disorder, bipolar disorder | 28 (20.4) | 252 (37.2) | 0 (0.0) | <0.01 | <0.01 | <0.01 |
| Mother's FD | 1 (0.7) | 27 (4.0) | 4 (1.2) | 0.21 | 1.00 | 0.04 |
| Mother's GORD | 14 (10.2) | 128 (18.9) | 14 (4.1) | 0.04 | 0.05 | <0.01 |
| Mother's IBD | 0 (0.0) | 5 (0.7) | 5 (1.5) | 0.31 | 0.15 | 0.26 |
| Mother's IBS | 12 (8.8) | 86 (12.7) | 24 (7.1) | 0.75 | 1.00 | 0.02 |
| Mother's migraine | 15 (10.9) | 230 (34.0) | 20 (5.9) | <0.01 | 0.24 | <0.01 |
| Mother's EDS, JHS | 0 (0.0) | 3 (0.4) | 1 (0.3) | 0.44 | 0.53 | 0.72 |
| Mother's obsessive-compulsive disorder | 2 (1.5) | 5 (0.7) | 3 (0.9) | 0.40 | 0.58 | 0.81 |
| Mother's intellectual disability | 0 (0.0) | 0 (0.0) | 0 (0.0) | N/A | N/A | N/A |
| Mother's schizophrenia | 1 (0.7) | 0 (0.0) | 1 (0.3) | 0.03 | 0.51 | 0.16 |

FD, functional dyspepsia; GORD, gastro-oesophageal reflux disease; IBD, inflammatory bowel disease; IBS, irritable bowel syndrome; EDS, Ehlers-Danlos syndrome; JHS, joint hypermobility syndrome

#### Phenotype 1

Most children in Phenotype 1 had allergic diseases (99.3%), and all mothers had allergic diseases (100%), which were significantly higher than in Phenotypes 2 and 3 (*p* < 0.01). In short, this phenotype was characterized as ‘AP with allergic predisposition’, suggesting relevance of allergic mechanisms in AP development in children.

#### Phenotype 2

In this phenotype, the frequency of allergic diseases is relatively high at 68.7%, but other comorbidities in the children were uncommon. In contrast, maternal comorbidities showed the highest frequencies in this phenotype: AP (70.0%), allergic diseases (40.0%), depressive and/or bipolar disorder (37.2%), GORD (18.9%), and migraine (34.0%). We termed Phenotype 2 as ‘AP with mothers’ comorbidities’.

#### Phenotype 3

The frequency of mother’s AP (46.2%) was the second highest in Phenotype 3. However, the frequencies of other comorbidities, including children’s allergic diseases and other maternal comorbidities were uncommon. This phenotype was termed ‘AP with the least comorbidities’.

#### Impact of allergic diseases and maternal comorbidities on paediatric AP

We further investigated the effect of allergic diseases and maternal comorbidity burdens on the frequency of paediatric AP. As the number of paediatric allergic diseases increased, the frequency of AP in children increased commensurately (Fig 3A). Specifically, 17.6% of children with ≥ 3 allergic diseases experienced AP, which was significantly more frequent than those with 0–2 allergic diseases (*p* < 0.01). Similarly, as the number of maternal comorbidities increased, the frequency of AP in children also increased (Fig 3B). In cases where mothers had ≥ 3 comorbidities, the frequency of paediatric AP was 25.6%, which was significantly higher than in cases with 0–2 comorbidities (*p* < 0.01). Furthermore, when comparing the high-risk children (those who met the criteria of having both ≥ 3 allergic diseases and ≥ 3 maternal comorbidities) to the others (control), the high-risk group had a significantly higher frequency of AP (*p* < 0.01) (Fig 3C).

**FIGURE 3.**
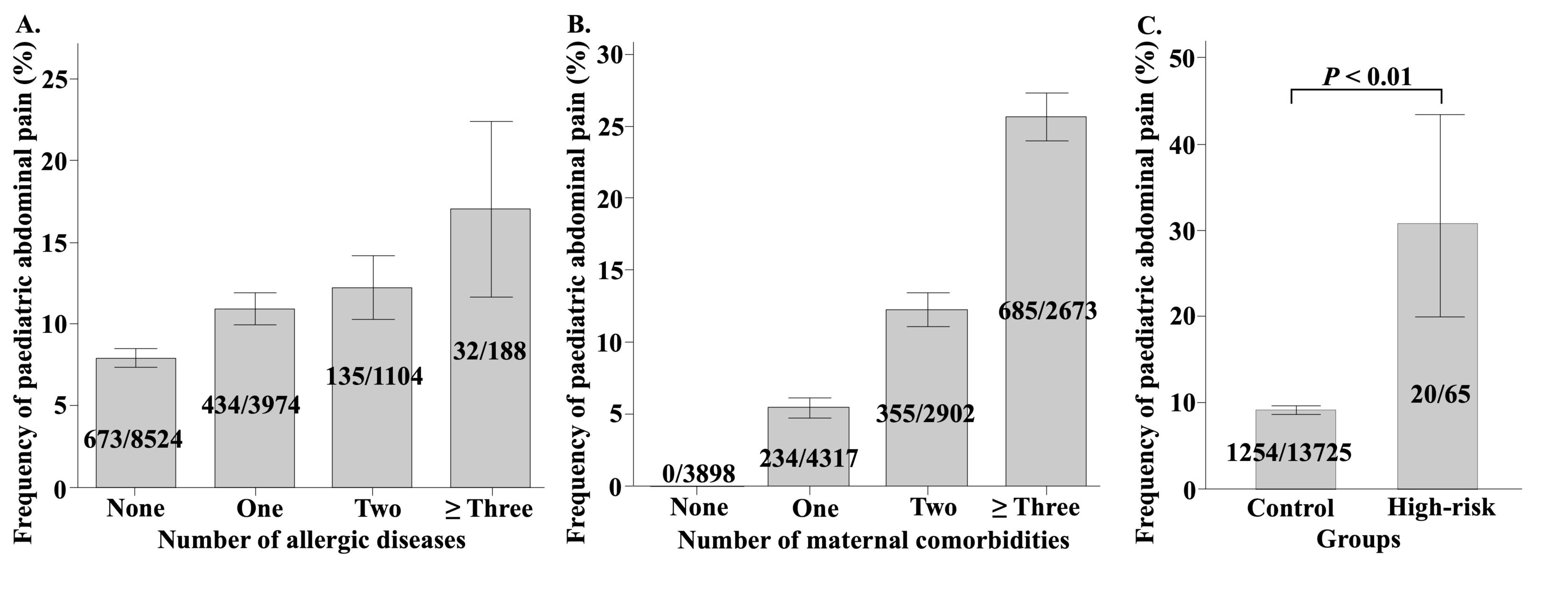
The frequency of paediatric abdominal pain depending on the number of allergic diseases and the number of maternal comorbidities A–B. The bar graphs illustrate the relationship between the frequency of paediatric abdominal pain (AP) and the number of allergic diseases in children, as well as the number of comorbidities in their mothers. Error bars in the figure represent the 95% confidence intervals (CI) for the frequency of paediatric AP. **C.** The high-risk group is defined as children with both ≥ 3 allergic diseases and ≥ 3 maternal comorbidities while the remaining children are regarded as the control group. The frequency of AP is compared between the high-risk and control groups.

### 3.3 Study 3. Development of ML predictive models for paediatric AP

Finally, we developed predictive models for paediatric AP. After evaluating several algorithms, CatBoost yielded the highest AUC of 0.67 (95% CI: 0.63–0.71) and was therefore selected for subsequent analyses (Fig 4A and S5 Table). The accuracy, sensitivity, specificity, PPV, and NPV of the CatBoost-based model were 0.62 (95% CI: 0.58–0.65), 0.68 (95% CI: 0.63–0.73), 0.55 (95% CI: 0.50–0.61), 0.62 (95% CI: 0.57–0.66), and 0.62 (95% CI: 0.57–0.68), respectively (Table 4).

**FIGURE 4.**
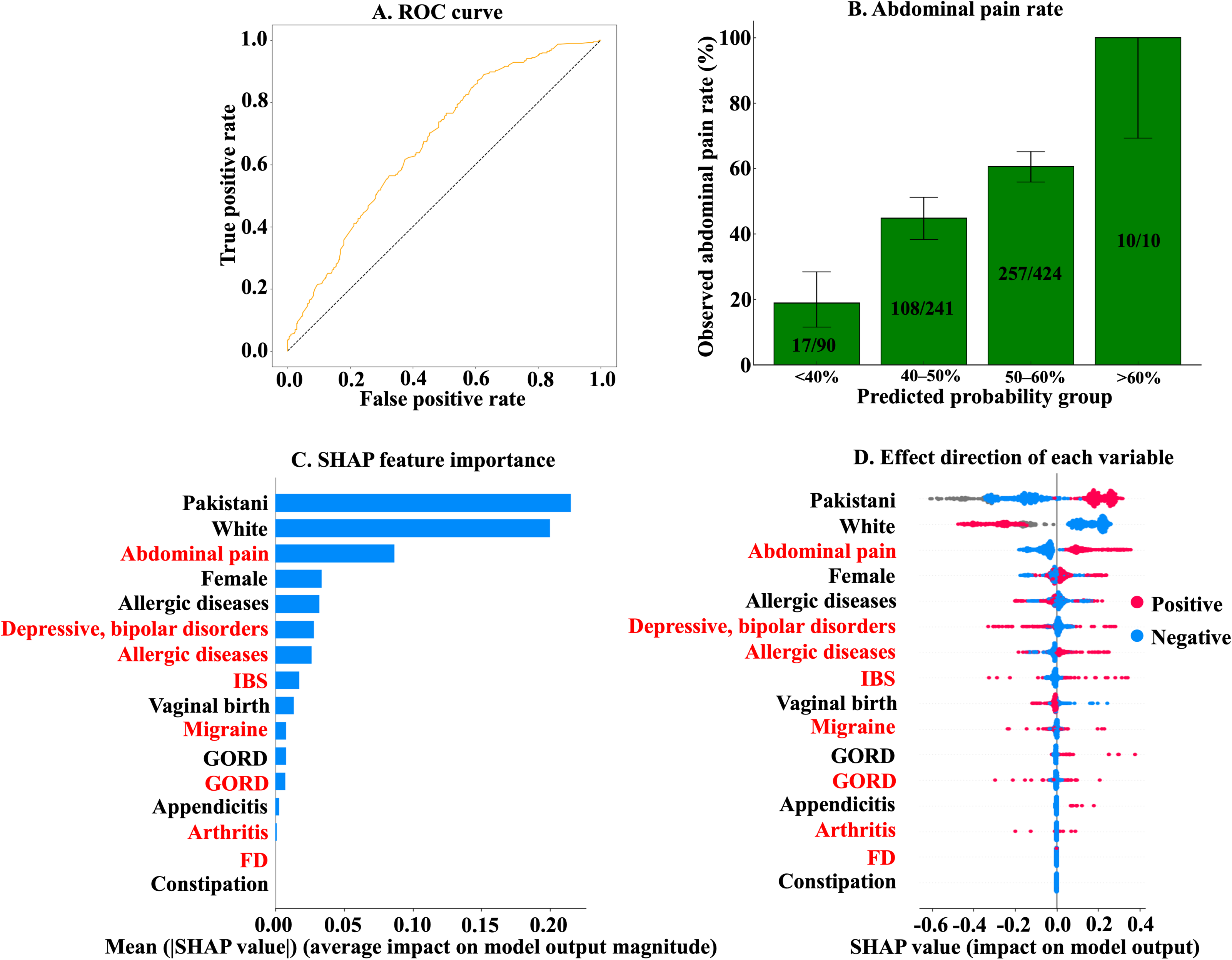
Results of supervised machine learning predictive models for paediatric abdominal pain. **A**. Receiver operating characteristic (ROC) curve of the CatBoost-based model. **B.** The y-axis represents the observed abdominal pain (AP) rate, while the x-axis indicates the predicted probability groups. Error bars represent the 95% confidence intervals of the observed AP rates. **C.** The mean Shapley Additive exPlanations (SHAP) value of each variable. Variables written in red represent maternal comorbidities, while those written in black represent variables related to the children themselves. **D.** The beeswarm plots show the SHAP feature importance and the direction of the effect of each variable on the model. The horizontal axis represents the SHAP value. Since all the variables used in the predictive models were binary, red and blue plots indicate positive or negative for each variable, respectively. For example, in the case of ‘Pakistani’, red plots tend to be distributed in the positive SHAP value and blue plots in the negative SHAP value. This indicates that Pakistani children are more likely to have AP.

**Table 4.** Results of predictive models for paediatric abdominal pain.

| Performance metrics | Outcomes |
| --- | --- |
| AUC (95% CI) | 0.67 (0.63–0.71) |
| Accuracy (95% CI) | 0.62 (0.58–0.65) |
| Sensitivity (95% CI) | 0.68 (0.63–0.73) |
| Specificity (95% CI) | 0.55 (0.50–0.61) |
| PPV (95% CI) | 0.62 (0.57–0.66) |
| NPV (95% CI) | 0.62 (0.57–0.68) |

Abbreviations: CI, confidence interval; PPV, positive predictive value; NPV, negative predictive value;
AUC, area under the curve

SHAP analysis indicated that ethnicity had a greater influence on the prediction than comorbidities, with White children being less likely and Pakistani children more likely to develop AP (Figs 4C and 4D). Maternal AP emerged as the most influential maternal comorbidity, while allergic diseases were the strongest predictor among paediatric comorbidities.

When stratified by the probability estimated by the CatBoost-based model, the observed rates of AP were 18.9%, 44.8%, 60.6%, and 100.0% in the <40%, 40–50%, 50–60%, and >60% probability groups, respectively (Fig 4B and Table 5). Compared to the lowest-probability group (<40%), the odds of AP were significantly higher in the 40–50% group (OR 3.49, 95% CI: 1.94–6.26, p < 0.01), 50–60% group (OR 6.61, 95% CI: 3.76–11.6, *p* < 0.01), and >60% group (OR could not be calculated, but *p* < 0.01).

**Table 5.**
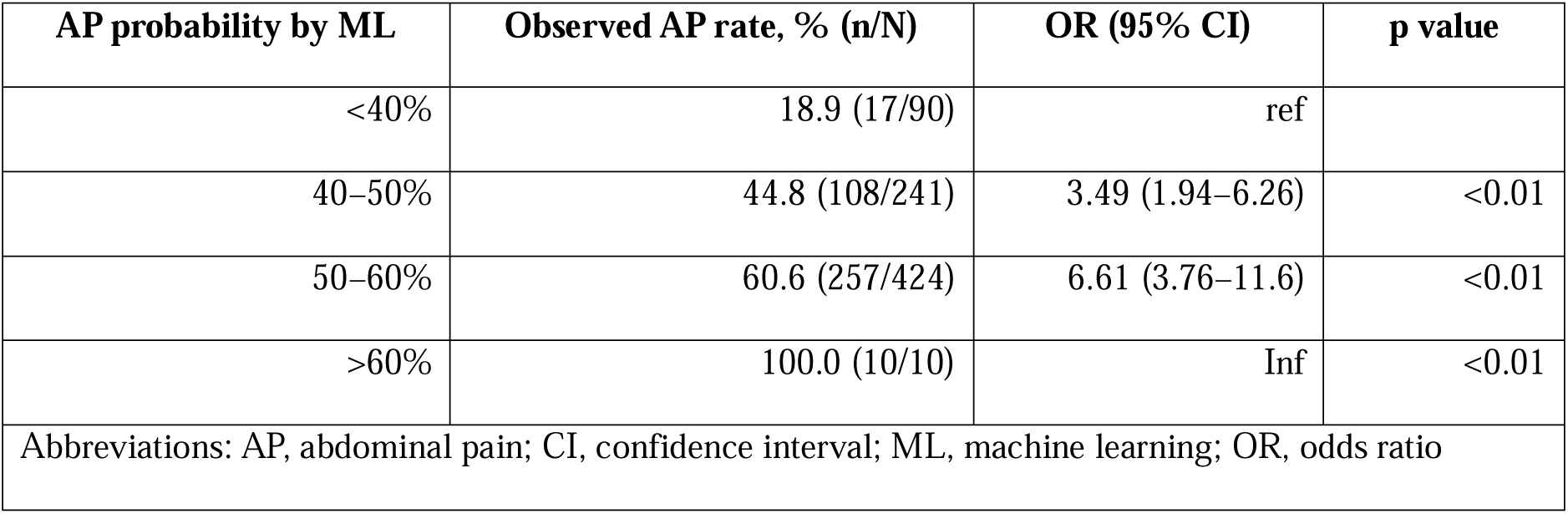
Risk stratification of paediatric abdominal pain based on the probability predicted by the CatBoost-based ML model.

## 4 Discussion

The prevalence of paediatric AP in our study cohort was 9.2 %. Several child and maternal factors, including paediatric ethnicity, GI and allergic diseases, as well as maternal AP, were associated with paediatric AP. ML-based clustering successfully identified three distinct phenotypes of paediatric AP, suggesting the presence of subgroups among children with AP. The characteristics of these phenotypes indicate that allergic diseases and maternal comorbidities are relevant to the development of AP in two of the three phenotypes. The predictive performance of the ML model was moderate when using the data from GP records. The most influential determinants of paediatric AP in the predictive model were child ethnicity, allergic predisposition, and maternal comorbidities, particularly maternal AP.

In this study, maternal GI comorbidities, such as AP and GORD, were significantly associated with paediatric AP. This finding is consistent with previous research showing that parental factors are related to functional AP in children [8,10,18]. For example, children of mothers with IBS or chronic pain tended to experience more GI symptoms or functional AP [18,25]. This association may be explained by parental influence on children’s pain experiences, whereby children may model their parents’ pain behaviours, and parents may reinforce their children’s pain complaints through solicitous responses [18,25–27]. Moreover, functional GI disorders, which are common causes of AP, are influenced by genetic factors [28], suggesting that the observed association may be partly due to shared genetic predisposition between mothers and children. Our findings underscore the important link between maternal factors and paediatric AP.

The ML clustering revealed three distinct phenotypes of paediatric AP. In all phenotypes, paediatric AP could not be simply explained by GI diseases. Phenotype 1 was characterized by the presence of allergic diseases. Some studies have reported that allergic diseases are significant risk factors for functional GI disorders such as IBS in children [9,17,29]. A recent study reported that IBS may be a food-induced disorder mediated by mast cell activation localized to the intestine [30]. Mast cells are known to be associated with visceral hypersensitivity [31]. These previous reports, along with our findings, imply that GI neuro-immune reactions could be one of the possible mechanisms underlying AP in children with allergic predispositions.

In Phenotype 2, where maternal comorbidities were associated, aforementioned parental mechanisms may play a role in the development of paediatric AP [18,25–28]. In contrast, in Phenotype 3, maternal AP was relatively common, occurring in 46.2% of cases. Therefore, some of the AP in children within Phenotype 3 may be explained by maternal influence. However, since the frequency of maternal comorbidities was much lower than in Phenotype 2, it is unlikely that all cases can be attributed to this factor. It is possible that the information available from GP records was insufficient to deeply phenotype children in Phenotype 3. Furthermore, 9.4% of children in this study were categorized as unclassified. With more detailed AP-related data, such as socioeconomic status, lifestyle, autonomic nervous system function, and gut microbiota composition, it may be possible to perform a deeper phenotypic analysis of children in Phenotype 3 and identify unclassified children as a distinct phenotype with specific characteristics [32–34].

The CatBoost-based model achieved a moderate AUC of 0.67 and identified the child’s ethnicity as a key factor, consistent with its significance in the logistic regression model. The BiB cohort is unique in that it primarily includes children of White (29.3%) and Pakistani ethnicities (39.5%) [15]. Socioeconomic status varies by ethnicity within the cohort [35], and lower socioeconomic status is a known risk factor for pain conditions in children, including AP [36]. These disparities may partly explain why ethnicity emerged as an important predictor.

Notably, approximately 60% of Pakistani couples represented in this cohort are reported to have consanguineous marriages, with 37% being first-cousin unions [39]. Children born from consanguineous marriages have been shown to have higher mortality rates and an increased number of primary care appointments compared to those born to non-consanguineous couples, indicating a higher burden of health issues [40]. Such consanguinity may also help explain why being of Pakistani ethnicity was a significant risk factor for paediatric AP in our model.

While the ML predictive model enabled a certain degree of risk stratification according to predicted probabilities, the substantial number of AP cases observed even in lower (40-50%) and intermediate (50-60%) probability groups suggests that the model’s discriminative power remains limited. This may be attributable to the fact that, although important associated factors such as allergic diseases and maternal AP were included as predictors in the ML model, variables directly reflecting the underlying pathophysiology of AP were not incorporated. Future models may benefit from incorporating variables that capture the underlying pathophysiology of AP, such as visceral hypersensitivity, altered gut-brain signalling, dysbiosis, and subclinical intestinal inflammation [41–43].

Our study is not without limitations. Firs, we were unable to distinguish between chronic and acute AP due to the lack of temporal detail in the available GP records. While this limits the precision of phenotype classification, it reflects real-world clinical practice where such distinctions are not always made or documented clearly. Moreover, the identified risk factors–such as allergic predisposition– are likely relevant to both types of AP, as supported by previous literature [17,44]. Most of our findings are also consistent with prior studies, lending support to the credibility and reliability of the data [40]. Therefore, despite this limitation, our findings provide valuable insights into the broader mechanisms of paediatric AP and remain relevant for understanding its development. Second, at the GP level, the exact cause of AP might not always be identifiable. Therefore, the criteria used for each diagnosis and the exact causes of AP in each case are unknown and was beyond our control. However, functional AP is likely the predominant cause, as previous community-based studies of AP in children suggest that approximately 80% of cases are classified as functional or non-medically explained, whereas organic diseases account for only 5–10% of cases [42]. Despite its complexity, real-world data of this nature holds significant potential for generating valuable insights, particularly when analysed using advanced methods such as ML [45]. Finally, although we evaluated the performance of our predictive models in an out-of-sample test partition, we did not fully validate them using a different cohort of children, which would further maximize generalizability. Moreover, a different cohort would also be useful for verifying the reproducibility of clustering. However, our study was an exploratory one and further research is now warranted to investigate the factors we have identified in detail.

In conclusion, our exploratory study identified paediatric and maternal comorbidities as significant associated factors for paediatric AP. Using data-driven clustering techniques, we uncovered three distinct phenotypes of paediatric AP. These findings provide a foundation for future research aimed at elucidating the precise mechanisms underlying paediatric AP. The predictive models developed in this study highlight the potential for early identification and intervention. Further studies incorporating more detailed data could refine phenotyping and predictive models, leading to more accurate predictions and paving the way for optimised, patient-personalised treatment strategies.

## Data Availability

All data produced in the present study are available upon reasonable request to the authors.

---

**S1 FIGURE.**
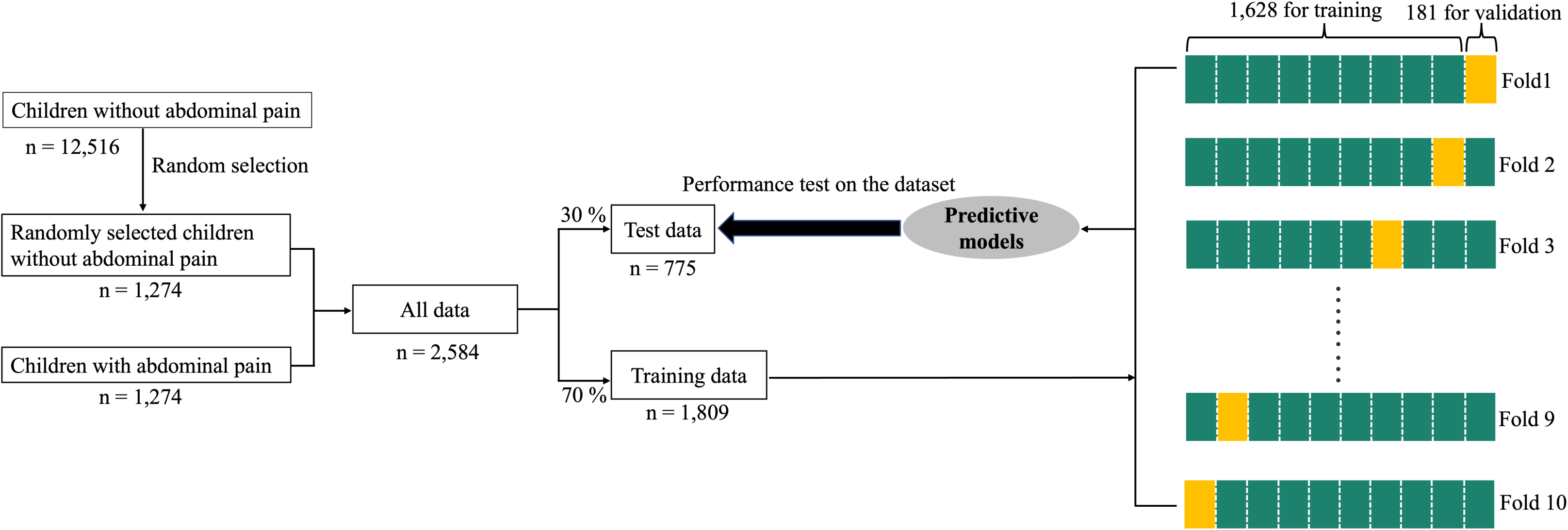
Workflow for developing predictive models. The entire dataset was randomly partitioned into 70% for model training and 30% for out-of-sample testing. Using the training data, we trained predictive models. Grid-search with 10-fold cross-validation was employed to tune the hyperparameters of each model using the training dataset. The training dataset was divided into 10 folds. Nine folds were used as the training dataset, and the remaining one-fold was used as the validation dataset. This process was repeated 10 times to optimize the hyperparameters of each predictive model. The performance of each model was evaluated using the test dataset.

