## Supplementary Tables for "An exploratory machine learning study on paediatric abdominal pain phenotyping and prediction"

**S1 Table. Investigated diseases and their corresponding systematized nomenclature of medicine clinical terms (SNOMED-CT) codes**

| **Disease name notation in this study** | **SNOMED-CT** | **SNOMED-CT code** |
| --- | --- | --- |
| Abdominal pain | Abdominal pain | 21522001 |
|  | Generalized abdominal pain | 102614006 |
|  | Abdominal wall pain | 162042000 |
| Allergic diseases | Asthma | 195967001 |
|  | Eczema | 43116000 |
|  | Allergic rhinitis caused by pollen | 21719001 |
|  | Urticaria | 126485001 |
| Appendicitis | Appendicitis | 74400008 |
| Arthritis | Multiple joint pain | 35678005 |
|  | Osteoarthritis | 396275006 |
|  | Lumbar spondylosis | 239880009 |
|  | Cervical osteoarthritis | 387800004 |
| Autism | Autistic disorder | 408856003 |
| Celiac disease | Celiac disease | 396331005 |
| Chronic fatigue syndrome | Chronic fatigue syndrome | 52702003 |
| Chronic muscle pain | Fibromyalgia | 203082005 |
| Constipation | Chronic constipation | 236069009 |
| Depressive disorder, bipolar disorder | Depressive disorder | 35489007 |
|  | Postpartum depression | 58703003 |
|  | Bipolar disorder | 13746004 |
| EDS, JHS | Generalized benign Joint hypermobility | 240261009 |
|  | Hypermobility syndrome | 85551004 |
|  | Ehlers-Danlos Syndrome | 398114001 |
|  | Hypermobile Ehlers-Danlos syndrome | 30652003 |
| FD | Nonulcer dyspepsia | 3696007 |
| GORD | Gastroesophageal reflux disease | 235595009 |
| IBD, colitis | Crohn's disease | 34000006 |
|  | Ulcerative colitis | 64766004 |
|  | Infectious colitis | 39341005 |
|  | Colitis presumed infectious | 79099006 |
|  | Diverticulitis of colon | 111359004 |
| IBS^*^ | Irritable bowel syndrome with diarrhea | 197125005 |
|  | Irritable bowel syndrome characterized by constipation | 440630006 |
|  | Irritable bowel syndrome characterized by alternating bowel habit | 440544005 |
|  | Irritable bowel syndrome | 10743008 |
| Intellectual disability | Intellectual disability | 110359009 |
| Migraine | Migraine | 37796009 |
| obsessive-compulsive disorder | Obsessive-compulsive disorder | 191736004 |
| Schizophrenia | Schizophrenia | 58214004 |
| ^*^IBS status was assessed only in mothers. Given that IBS is defined by the presence of abdominal pain, its inclusion in this study—designed to investigate factors associated with abdominal pain—was considered methodologically inappropriate.  FD, functional dyspepsia; GORD, gastro-oesophageal reflux disease; IBD, inflammatory bowel disease; IBS, irritable bowel syndrome; EDS, Ehlers-Danlos syndrome; JHS, joint hypermobility syndrome; SNOMED-CT, Systematized Nomenclature of Medicine Clinical Terms | | |

**S2 Table. Frequency of variables used in machine learning predictive models**

|  | **Children without abdominal pain**  **(n = 1,274)** | **Children with abdominal pain**  **(n = 1,274)** | ***p* value** |
| --- | --- | --- | --- |
| Female | 590 (46.3) | 659 (51.7) | 0.01 |
| Route of birth, vaginal | 941 (73.9) | 943 (74.0) | 0.96 |
| Pakistani | 485 (45.8) | 703 (58.6) | <0.01 |
| White British | 382 (36.1) | 267 (22.3) | <0.01 |
| Other ethnicities | 191 (18.1) | 229 (19.1) | 0.56 |
| Allergic diseases | 457 (35.9) | 482 (37.8) | 0.32 |
| Appendicitis | 1 (0.1) | 19 (1.5) | <0.01 |
| Constipation | 13 (1.0) | 31 (2.4) | 0.01 |
| GORD | 35 (2.7) | 62 (4.9) | 0.01 |
| Mother’s abdominal pain | 392 (30.8) | 557 (43.7) | <0.01 |
| Mother’s allergic disease | 492 (38.6) | 515 (40.4) | 0.37 |
| Mother’s arthritis | 12 (0.9) | 15 (1.2) | 0.7 |
| Mother’s depressive disorder, bipolar disorder | 274 (21.5) | 252 (19.8) | 0.3 |
| Mother’s FD | 14 (1.1) | 20 (1.6) | 0.39 |
| Mother’s GORD | 83 (6.5) | 102 (8.0) | 0.17 |
| Mother’s IBS | 85 (6.7) | 101 (7.9) | 0.25 |
| Mother’s migraine | 187 (14.7) | 222 (17.4) | 0.07 |
| FD, functional dyspepsia; GORD, gastro-oesophageal reflux disease; IBD, inflammatory bowel disease; IBS, irritable bowel syndrome | | | |

**S3 Table. Characteristics of all children (N = 13,790)**

| **Background** |  |
| --- | --- |
| Average age at the extraction ± SD (years) | 8.3 ± 1.1 |
| Average age at the diagnosis of abdominal pain ± SD (years) | 5.6 ± 2.7 |
| Gender, female, n (%) | 6,674 (48.4) |
| Route of birth, (vaginal)*, n (%) | 10,358 (76.8) |
| Ethnicity**, n (%) |  |
| Pakistani | 5,453 (47.1) |
| White British | 4,038 (34.9) |
| Other ethnicities | 2,076 (17.9) |
| Abdominal pain, n (%) | 1,274 (9.2) |
| **Frequency of diseases, n (%)** |  |
| Allergic diseases | 5,266 (38.2) |
| Asthma | 1,383 (10.0) |
| Hay fever | 851 (6.2) |
| Urticaria | 695 (5.0) |
| Eczema | 3,828 (27.8) |
| Appendicitis | 51(0.4) |
| Arthritis | 7 (0.1) |
| Celiac disease | 33 (0.2) |
| Constipation | 199 (1.4) |
| FD | 7 (0.1) |
| GORD | 408 (3.0) |
| IBD, colitis | 6 (0.04) |
| Migraine | 69 (0.5) |
| EDS, JHS | 13 (0.1) |
| Autism | 40 (0.3) |
| Intellectual disability | 18 (0.1) |
| Mother’s abdominal pain | 4,616 (33.5) |
| Mother’s allergic disease | 5,540 (40.2) |
| Mother’s appendicitis | 107 (0.8) |
| Mother’s arthritis | 604 (4.4) |
| Mother’s Celiac disease | 87 (0.6) |
| Mother’s chronic fatigue syndrome | 28 (0.2) |
| Mother’s chronic muscle pain | 197 (1.4) |
| Mother’s constipation | 138 (1.0) |
| Mother’s depressive disorder, bipolar disorder | 2,941 (21.3) |
| Mother’s FD | 195 (1.4) |
| Mother’s GORD | 1,047 (7.6) |
| Mother’s IBD | 94 (0.7) |
| Mother’s IBS | 1,003 (7.3) |
| Mother’s migraine | 2,130 (15.4) |
| Mother’s EDS, JHS | 52 (0.4) |
| Mother’s obsessive-compulsive disorder | 71 (0.5) |
| Mother’s schizophrenia | 8 (0.1) |
| Mother’s intellectual disability | 45 (0.3) |
| * 311 values are missing in route of birth. ** 2223 values are missing in ethnicity. | |
| SD, standard deviation; FD, functional dyspepsia; GORD, gastro-oesophageal reflux disease; IBD, inflammatory bowel disease; IBS, irritable bowel syndrome; EDS, Ehlers-Danlos syndrome; JHS, joint hypermobility syndrome | |

**S4 Table. Non–significant results of logistic regression analysis for the diagnosis of paediatric abdominal pain**

|  | **OR (95% CI)** | ***p* value** |
| --- | --- | --- |
| Gender, female | 0.94 (0.82-1.08) | 0.40 |
| Route of birth, vaginal (vs. caesarean) | 0.93 (0.80-1.09) | 0.36 |
| Arthritis | 21000 (0.00–Inf) | 0.95 |
| FD | 4.01 (0.73-21.90) | 0.11 |
| IBD | 4.00 (0.36-44.20) | 0.26 |
| Autism | 0.89 (0.27-2.89) | 0.84 |
| Intellectual disability | 1.33 (0.22-7.99) | 0.75 |
| EDS, JHS | 8.02 (0.90-71.90) | 0.06 |
| Mother’s appendicitis | 1.87 (0.88-3.98) | 0.11 |
| Mother’s constipation | 1.00 (0.25-4.01) | 1.00 |
| Mother’s Celiac disease | 1.54 (0.74-3.17) | 0.25 |
| Mother’s depressive disorder, bipolar disorder | 1.11 (0.94-1.30) | 0.22 |
| Mother’s IBD | 0.55 (0.27-1.12) | 0.10 |
| Mother’s EDS, JHS | 1.50 (0.63-3.58) | 0.36 |
| Mother’s obsessive-compulsive disorder | 1.00 (0.34-2.93) | 1.00 |
| Mother’s schizophrenia | 1.54 (0.68-3.53) | 0.30 |
| Mother’s intellectual disability | 2.00 (0.13-32.00) | 0.62 |
| IBD, inflammatory bowel disease; EDS, Ehlers-Danlos syndrome; JHS, joint hypermobility syndrome | | |

**S5 Table. Results of predictive models for paediatric abdominal pain using various machine learning algorithms**

|  | **AUC (95% CI)** | **Accuracy (95% CI)** | **Sensitivity (95% CI)** | **Specificity (95% CI)** | **PPV (95% CI)** | **NPV (95% CI)** |
| --- | --- | --- | --- | --- | --- | --- |
| XGBoost | 0.63 (0.59–0.67) | 0.59 (0.56–0.62) | 0.61 (0.56–0.62) | 0.57 (0.52–0.62) | 0.60 (0.55–0.65) | 0.58 (0.53–0.63) |
| Random Forest | 0.63 (0.59–0.67) | 0.58 (0.55–0.62) | 0.58 (0.53–0.62) | 0.59 (0.54–0.64) | 0.60 (0.55–0.65) | 0.57 (0.52–0.62) |
| CatBoost | 0.67 (0.63–0.71) | 0.62 (0.58–0.65) | 0.68 (0.63–0.73) | 0.55 (0.50–0.61) | 0.62 (0.57–0.66) | 0.62 (0.57–0.68) |
| LightGBM | 0.63 (0.59–0.67) | 0.58 (0.55–0.62) | 0.61 (0.57–0.66) | 0.55 (0.50–0.60) | 0.59 (0.54–0.63) | 0.57 (0.52–0.62) |
| Abbreviations: XGBoost, eXtreme Gradient Boosting; CatBoost, Categorical Boosting; LightGBM, Light Gradient Boosting Machine; CI, confidence interval; PPV, positive predictive value; NPV, negative predictive value; AUC, area under the curve | | | | | | |
